# Investigating the shared genetic architecture between type 2 diabetes and stroke

**DOI:** 10.1101/2025.03.24.25324577

**Authors:** Tania Islam, Jian Zeng, Mohammad Ali Moni

## Abstract

Individuals with type 2 diabetes (T2D) have an approximately two-fold higher risk of stroke compared to those without diabetes. However, the genetic relationships and biological mechanisms underlying T2D and stroke remain incompletely understood. This study aims to explore the shared genetic architecture and causal relationships of T2D and stroke using large-scale genome-wide association study (GWAS) datasets. We observed a significant causal genetic overlap (∼900 variants) and global genetic correlation between T2D and stroke (rG = 0.35, P = 3.5 × 10^−24^). We identified a significant genetic causal association of T2D, independent from its confounders such as coronary artery disease, BMI, and educational attainment, on the stroke risk. This causal association was not affected by reverse causations. A cross-trait meta-analysis and functional annotation revealed 76 independent SNPs, of which 19 were lead SNPs with genome-wide significance (*P*-value < 5.8×10-08), shared between T2D and stroke, among them 10 independent lead SNPs are novel. Summary-based Mendelian randomization (SMR) identified 109 putative causal genes for T2D and 25 for stroke, respectively, after multiple corrections (Bonferroni *P*-value < 5.23×10-06; *P*_HEIDI_ > 0.01) where seven genes were found overlapping between T2D and stroke; of which *SREBF1, LTBP3, FAM234A, PABPC4*, and *RMC1* were novel for stroke. Pathway-based functional enrichment analyses identified critical pathways, including regulation of autophagy, negative regulation of insulin secretion, and positive regulation of cholesterol biosynthetic process, as shared molecular targets. Overall, the findings of this study provide novel risk loci, causal genes, and pathways shared between T2D and stroke, suggesting molecular targets for these co-occurring diseases.

## Introduction

Stroke is a complex neurological disorder, resulting in approximately 12% of deaths and 6% of disability-adjusted life years around the world [1, 2]. Type 2 diabetes is a metabolic disease characterized by insufficient insulin secretion, resulting in substantial changes in the body’s glucose homeostasis [3], affecting around 10.5% of adults aged 20 to 79 [4]. Alongside the effect of aging, different genetic and environmental factors contribute to T2D development, and T2D has been identified as a risk factor for ischemic stroke and cardiovascular diseases, including coronary artery disease (CAD) [5, 6]. Observational epidemiological studies have suggested that T2D and its risk variables such as body mass index (BMI), insulin resistance, and hyperglycemia are associated with stroke risk [7–9]. Individuals with diabetes have an approximately two-fold higher risk of stroke compared to those without diabetes, experience worse post-stroke complications, and have a higher likelihood of stroke recurrence [7, 10–15]. However, the extent to which T2D has a causal effect on stroke risk remains unclear due to shared genetic factors and potential confounders [16, 17].. In particular, cardiovascular conditions such as coronary artery disease (CAD) may mediate or explain part of this relationship. Therefore, in this study, we have investigated whether T2D has a direct causal effect on stroke after accounting for CAD. In addition, we also include BMI and educational attainment (EA) as potential confounders to determine the independent contribution of T2D on stroke risk [16, 17].

In this study, leveraging the largest available GWAS datasets, we have investigated the shared polygenic overlap, genetic correlation, causal relationships, and risk genes between T2D and stroke. We further performed cross-trait GWAS meta-analysis identified novel SNPs associated with T2D and stroke. Gene-based analysis was conducted to identify functional genes, and pathway-based analysis revealed distinct biological mechanisms shared between T2D and stroke.

## Methods

### Study design

Figure 1 depicts the core premise of the present study. We conducted three extensive analyses at the SNP, gene, and functional enrichment levels.

**Figure 1.**
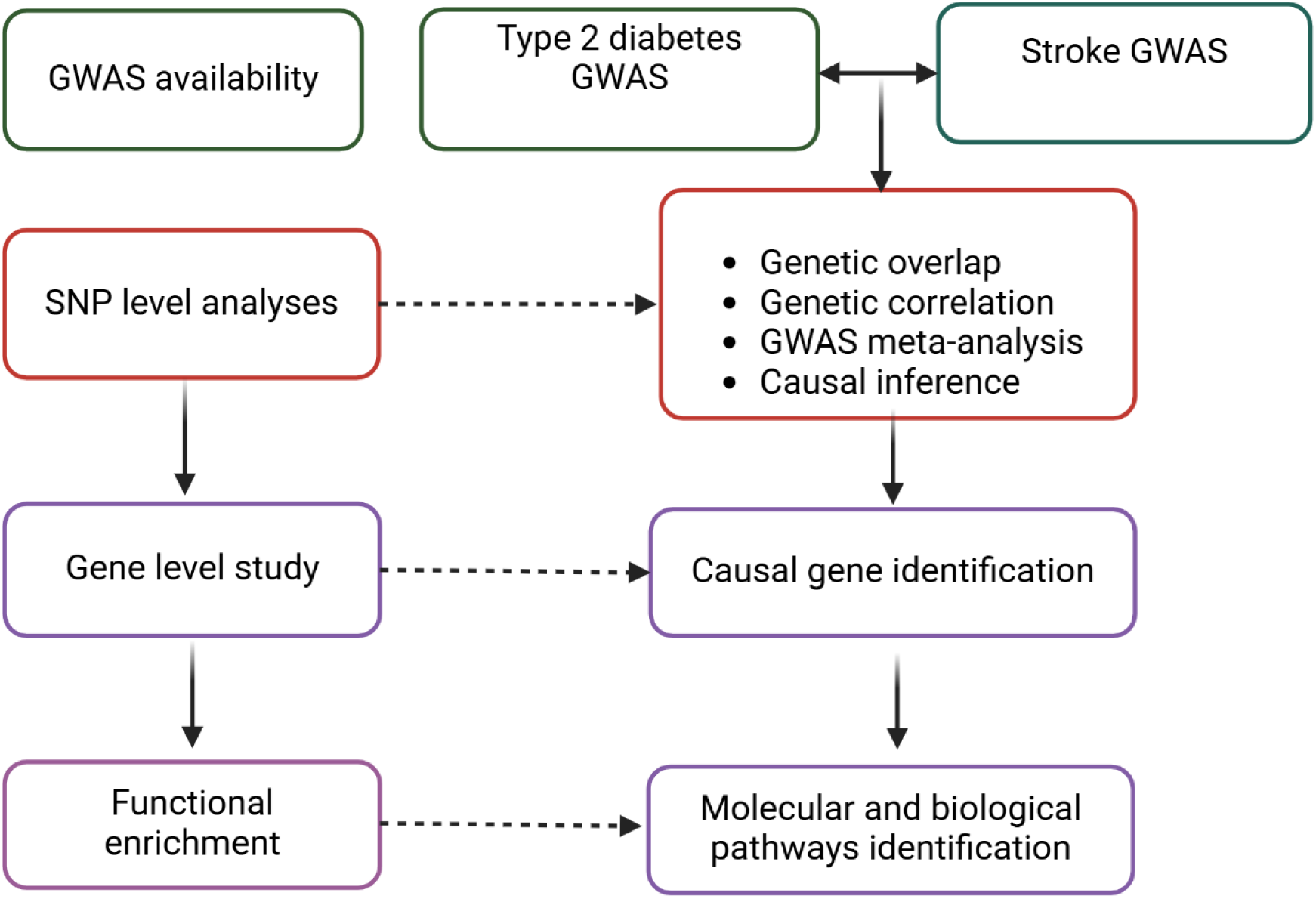
The schematic diagram represents the overall study design. First, SNP-level analyses were performed to understand shared genetic etiology, genetic correlation, causal relationship analysis, and multi-trait GWAS meta-analysis between T2D and stroke. Second, we did gene-level analysis to explore potential causal risk genes between T2D and stroke. Finally, we performed functional enrichment analysis to identify important pathways shared between T2D and stroke. This figure was created in BioRender.com.

Firstly, we performed SNP level analysis to characterized shared genetic architecture at SNP level. We used MiXeR software to investigate the overlapping polygenic architecture of T2D and stroke. We also performed an LDSC analysis to determine the genetic correlation between them. Furthermore, a cross-trait meta-analysis was conducted by integrating GWAS datasets for T2D and stroke to identify SNPs and risk loci that overlapped in T2D and stroke. Further, conditional and joint analysis was conducted to avoid the effects of covariates of T2D and stroke. Subsequently, we used Mendelian randomization (MR) to determine potential causal associations between T2D and stroke. Second, we then performed gene-based analyses utilizing the summary-data-based Mendelian randomization method to identify potential causal genes shared by T2D and stroke. Finally, we performed functional enrichment analyses to understand the critical biological routes of shared between T2D and stroke.

### Description of GWAS datasets

The DIAGRAM consortium (https://www.diagram-consortium.org/) provided T2D GWAS summary data (accessed on 1^st^ September 2024) which contains European ancestry 242283 cases, 1569734 controls of T2D GWAS [18]. We also obtained stroke GWAS summary data from the GIGASTROKE consortium available in the GWAS Catalogue (GCST90104539), which contains 73,652 stroke cases, and 1,234,808 healthy individuals in European ancestry controls (sTable 4.1) [19].

### Gaussian causal mixture modelling method (MiXeR)

To estimate the polygenic overlap between T2D and stroke, we performed MiXeR model (MiXeR v1.3) using respective GWAS summary data. We performed univariate analysis to estimate the number of trait-influencing loci for T2D and stroke, respectively. Next, bivariate analysis was performed estimate the genome-wide genetic overlap. The MiXeR framework models the genetic effects as a mix of four type of SNPs: (i) SNP that do not affect T2D and stroke, (ii) SNPs only affect T2D, (iii) SNPs only affect stroke, (iv) SNPs affect both T2D and stroke. The Venn diagram represents the distribution of shared and unique SNPs of T2D and stroke. The details of MiXeR methods can be found in the original publication [20] and code are available in online (https://github.com/precimed/mixer).

### Global genetic correlation assessment between T2D and stroke

To estimate the heritability and genetic correlation between T2D and stroke, we performed linkage disequilibrium score regression (LDSC) analysis [21] in (LDSC, v1.0.1, https://github.com/bulik/ldsc) software. LDSC estimates and distinguishes inflated test statistics from confounding biases, the contributions of polygenicity, sample overlap, and population stratification to the heritability and genome-wide genetic correlation among traits [21]. Firstly, we calculated SNP-based heritability (*h*^2^_SNP_) for T2D and stroke, respectively. Secondly, we analyzed bivariate LDSC to explore the cross-trait genetic correlation (r_g_) between T2D and stroke.

### Cross-trait GWAS meta-analysis

To identify novel shared genetic variants associated with both T2D and stroke, we used the multi-trait analysis of GWAS (MTAG) method [22] (version 1.0.8). MTAG is powerful approach because it can handle potential sample overlap between the GWAS datasets and maximize the statistical power to detect shared genetic variants. In this study, we utilized GWAS summary statistics for T2D and stroke. The GWAS summary data were pre-processed and quality-controlled before running MTAG. This involved filtering SNPs based on p-values, minor allele frequency (MAF), and linkage disequilibrium (LD), with a threshold of minor allele frequency ≤ 0.01 and removing any strand-ambiguous SNPs. In addition, SNPs with missing or invalid p-values, effect sizes, or standard errors were excluded. After quality control, 5,906,777 SNPs were retained for T2D, and 7,431,364 SNPs were retained for stroke. Strand ambiguous SNPs were removed to ensure accuracy, and after merging the datasets, 3,351,549 SNPs remained for MTAG analysis.

MTAG models the joint distribution of summary statistics across stroke and T2D using Z-scores of each SNP. It accounts for differences in SNP heritability across traits and estimates the upper bound of the false discovery rate (maxFDR) to ensure reliable results. GWAS summary statistics were processed through the MTAG pipeline, which estimated Omega (covariance of genetic effect sizes) to account for shared genetic architecture. The output included GWAS and MTAG mean chi-squared statistics, along with estimates of Omega and Sigma (variance-covariance matrix of genetic effects) [22]. SNPs surpassing the genome-wide significance threshold (*p* < 5×10^−8^) were retained for further analysis.

### Genomic loci characterization and independent SNP identification

To identify the potential independent lead SNPs linked to T2D and stroke (overlapped SNPs), we used FUMA (functional mapping and annotation) software (https://fuma.ctglab.nl) [23]. We used the meta-analysis results of T2D and stroke as input for FUMA. FUMA uses LD clumping to identify independent SNPs with *r*^2^ < 0.6 and independent lead SNPs without LD (*r*^2^ < 0.1). The physical regions within 250kb of lead SNP were identified as genomic loci. The lead SNPs within this region were consolidated into a single locus. The 1000 Genomes Phase 3 (EUR) dataset was utilized to estimate the LD information. Finally, the GWAS catalog was searched to identify novel independent lead SNPs for T2D and stroke with genome-wide significant levels to identify whether they had been previously reported as genome-wide significant.

### Multi-trait conditional analysis

We performed a multi-trait-based conditional and joint analysis (mtCOJO) as described by Zhu et al. (2018), [24], which was implemented in GCTA software (http://cnsgenomics.com/software/gcta/#mtCOJO), to estimate the effect of T2D on stroke while accounting for genetic effects of other covariates. First, we conditioned the GWAS summary data for T2D on CAD to obtain T2D SNP effects independent of CAD’s genetic influence. Next, we extended the mtCOJO model to jointly condition T2D on CAD, body mass index (BMI), and educational attainment (EA). The conditioned T2D SNP effects from these steps were used in Mendelian randomization (GSMR) analysis to estimate the causal effect of T2D on stroke.

### Causal relationship assessment using Mendelian randomization studies

To assess causal relationships between T2D and stroke, we conducted bidirectional two-sample Mendelian randomization (2SMR) analyses [25]. We used the R statistical package (https://mrcieu.github.io/TwoSampleMR/) to execute 2SMR analyses. In the forward MR analysis, we estimated the causal effect of T2D on stroke using three exposure scenarios: (i) unadjusted T2D; (ii) T2D conditioned on CAD, and (iii) T2D conditioned on CAD, BMI, and EA. Stroke GWAS data served as the outcome. In the reverse direction, we assessed the causal effect of stroke on T2D using: (i) unadjusted stroke, (ii) stroke conditioned on CAD, and (iii) stroke conditioned on CAD, BMI, and EA, with T2D as the outcome.

For both studies, independent (LD clumping at *r*^2^ < 0.001) SPNs with genome-wide significant levels (p<5×10^−8^) were used as instrumental variables (IVs). We also considered the F-statistics value >20 (calculated as *β^2^/SE^2^*) for each IVs to ensure IVs strength. The inverse variance weighted (IVW) method [26] investigated the causal relationship between T2D and stroke and vice versa. Other MR techniques, such as the MR-Egger, the weighted median [32], and the ’Mendelian randomization pleiotropy residual sum and outlier’ (MR-PRESSO) [33], were also used to estimate the causal genetic relationship between traits. We also performed a sensitivity analysis to identify the heterogeneity of the effects of SNPs associated with stroke; the I^2^ index and Cochran’s Q statistic were used for MR-IVW analysis, while the Rucker’s Q statistic was used for MR-Egger analysis, and p < 0.05 indicating the presence of heterogeneity [27]. We used the MR-Egger technique to see how much directional pleiotropy influences risk estimates from intercept testing, and p > 0.05 suggests the absence of pleiotropy [28]. As a complementary approach, we performed a generalized summary data-based Mendelian randomization (GSMR) analysis to estimate the causal effect of exposure (T2D) on the outcome (stroke) [24]. Briefly, GSMR eliminates the SNPs with multiple alleles or missing values and minor allele frequency (MAF) > 0.01. Next, GSMR employs clumping analysis to keep the independent genome-wide SNPs between exposure (T2D) and outcome (stroke) with genome-wide significant SNPs (P < 5×10^−8^ and r^2^ <0.01). The heterogeneity in dependent instruments (HEIDI) outlier test was performed to exclude the pleiotropic SNPs to capture the causal influence of the instrumental variable of exposure on the outcome.

### Gene-based analysis

We used MTAG-generated T2D and stroke GWAS data to perform summary-based Mendelian randomization (SMR) analyses; the SMR method is explained in more detail in the original paper by Zhu and colleagues [29]. SMR separately integrates GWAS summary data with eQTL data to identify causal genes. In SMR, genetic variants (SNPs) serve as instrumental variables (IVs) to estimate the effect of gene expression on disease phenotypes. The method leverages summary statistics from GWAS (for the disease phenotype) and eQTL studies (for gene expression). To identify the causal relationship, we calculate the SMR effect estimate, which is the ratio of the SNP’s effect on the phenotype (bzy) to its impact on the exposure (bzx). To differentiate pleiotropy from linkage, we applied the HEIDI test, which assesses whether a single causal variant explains the exposure and the outcome. Associations with PHEIDI < 0.01 were excluded as they suggest linkage rather than pleiotropy. We set the threshold for significance at PHEIDI > 0.01, and Bonferroni corrected *P*-value < 5.02×10^−6^ to identify pleiotropic or causal genes of interest.

### Gene set enrichment analysis

We performed functional enrichment analysis of common genes of T2D and stroke using Enrichr bioinformatics tools, accessed on 25^th^ September 2024 [30]. Enrichr is the most significant functional enrichment database for exploring gene ontology terms and pathways [30]. Adjusted p-values < 0.05 (Benjamini-Hochberg correction) and combined scores were used to determine statistical significance. Significant pathways were further explored in the context of their roles in metabolic and vascular processes.

## Results

### Univariate and Polygenic overlap and genetic correlation analysis

Univariate analysis using MiXerR showed substantial polygenicity of both T2D and stroke, with T2D exhibiting higher polygenicity compared to stroke. Bivariate modeling suggests substantial polygenic overlap between T2D and stroke, with sufficient quality of model fit based on log-likelihood plots and Akaike information criteria (Figure 2). Approximately 900 putatively causal SNPs were identified as overlapping between T2D and stroke.

**Figure 2.**
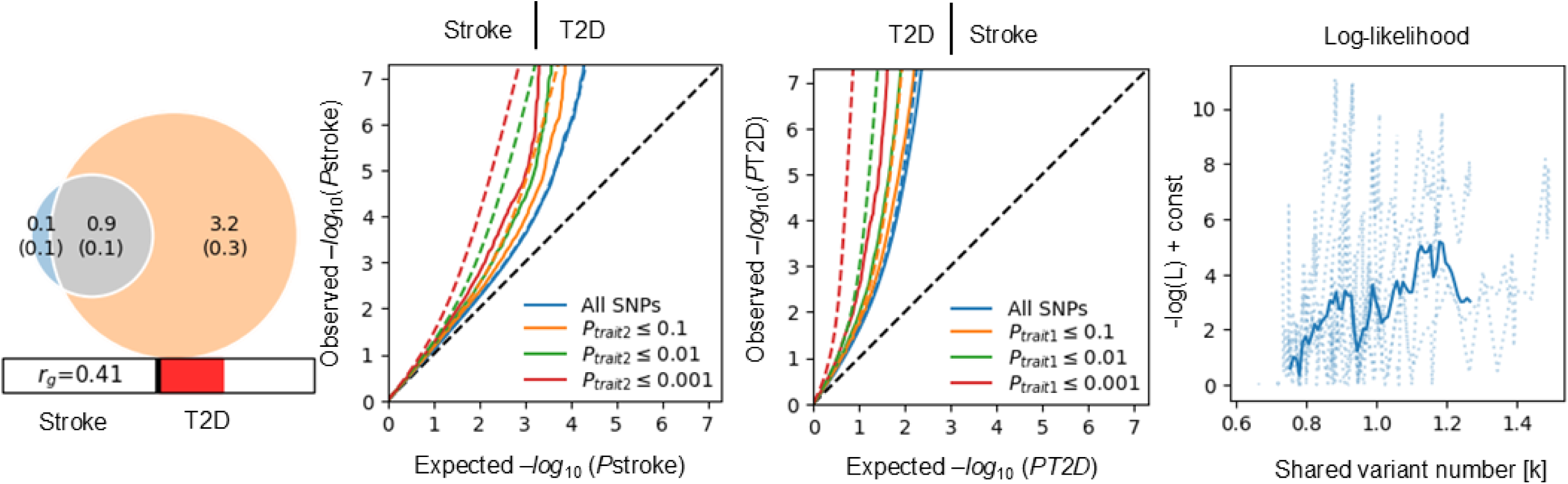
Venn diagram represents unique and shared polygenic elements at the causal level. The grey color indicates polygenic overlap between stroke (blue) and T2D (orange). The number represents the estimated causal variants (in thousands) explaining 90% of SNP heritability in each phenotype, followed by the standard error. Circle size shows the degree of polygenicity and rg=0.41 (MiXeR) demonstrates the genetic correlation between stroke and T2D. Conditional Q–Q plots depict observed versus expected −log_10_ p-values for stroke (left) as a function of significance of association with a T2D at the level of p ≤ 0.1 (orange lines), p ≤ 0.01 (green lines), p ≤ 0.001 (red lines). The blue line shows all SNPs. Dotted lines in blue, orange, green, and red indicate model predictions for each stratum. The black dotted line represents null expectation. log-likelihood as a function of the shared variant number (in thousands, [k]) between stroke and T2D.

We observed that SNP-based liability scale heritability (h2SNP) for T2D is 0.316 (standard error, SE=0.0193), suggesting that 31.6 % of the variation in disease risk is attributed to the common risk variants associated with T2D. In parallel, we observed that SNP-based liability scale heritability (h2SNP) for stroke is 0.02 (0.0023), indicating that 2% of the variation in disease risk is attributed to common genetic variants associated with stroke (Table 1). LDSC genetic correlations analysis demonstrated a significant global genetic correlation between T2D and stroke (rG = 0.35, P = 3.5 × 10^−24^). SNP-based heritability estimates and the genetic correlation between T2D and stroke are presented in Table 1.

**Table 1.**
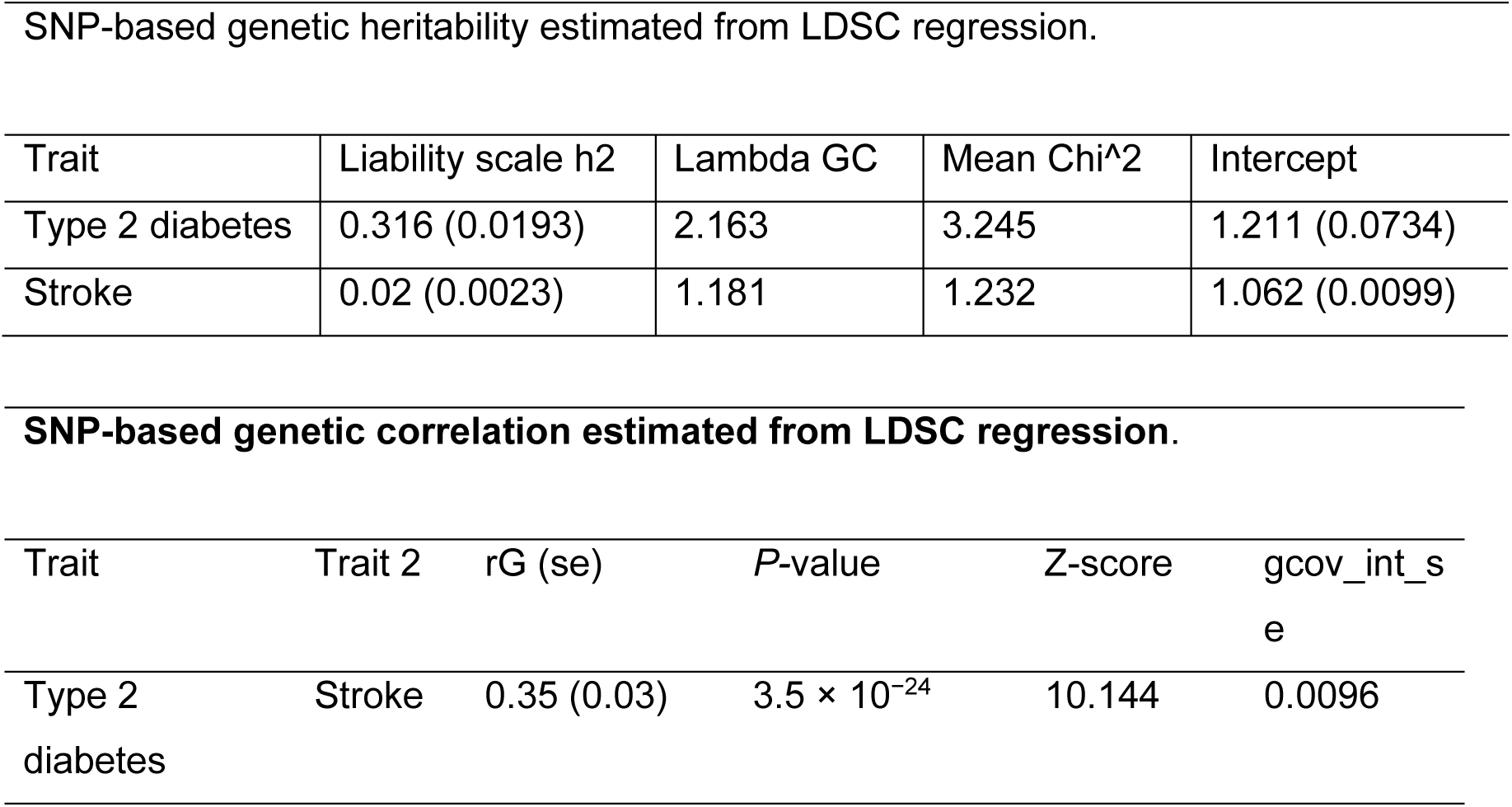
Estimating heritability and genetic correlation of type 2 diabetes and stroke.

### Identification of overlapping risk SNPs

To identify novel SNPs and loci that were not genome-wide significant in individual genome-wide association studies (GWAS) for type 2 diabetes (T2D) and stroke (5 × 10−⁸ < P_GWAS < 0.05) but became significant following cross-trait meta-analysis (P_meta-analysis_ < 5 × 10−⁸). Thus, we leveraged multi-trait analysis of GWAS (MTAG)-derived summary statistics for T2D and stroke as input for functional mapping and annotation (FUMA) analysis. Specifically, FUMA identified 848 lead-independent SNPs ((linkage disequilibrium, r² < 0.1) associated with T2D and 73 lead-independent SNPs r² < 0.1) associated with stroke (sTable 2 - sTable 3) respectively. Among these independent lead SNPs, 19 were shared between T2D and stroke (sTable 4), of which 10 were classified as novel according to GWAS Catalog. These novel SNPs, namely rs180726800, rs140908036, rs116369954, rs10885410, rs4918791, rs10750766, rs10885414, rs35801464, rs1016287, rs140820620, are jointly implicated in T2D and stroke pathogenesis (sTable 5).

### Mendelian randomization analysis to assess causal relationships Before conditional analysis MR results

To identify the potential causal genetic relationships between T2D and stroke, we performed two samples of Mendelian randomization (MR) analyses. We identified 460 independent genome-wide significant SNPs from T2D as instrumental variables (IVs) to determine the causal association of T2D (exposure) on stroke (outcome) (sTable 6). MR analysis supports evidence for the putative causal genetic influence of T2D on stroke risk, as indicated by the IVW method (OR = 1.11, 95% CI: 1.06-1.13, *P* = 1.33 x 10^-21^). The IVW method finding was consistent across sensitivity analyses, including weighted median (OR = 1.09, 95% CI: 1.06-1.122, *P* = 7.91×10^-09^), MR-Egger (OR= 1.09, 95% CI: 1.05-1.14, *P* = 7.8 x 10^-5^) (Table 2). In addition, the recently developed GSMR method, which accounts for LD and pleiotropy, further supported a significant genetic association between T2D and stroke risk (*P* = 1.27 ×10^-43^).

**Table 2.** Bidirectional causal genetic relationship between type 2 diabetes and stroke using MR analysis.

| Exposure | Outcome | IVW |  | Weighted median |  | MR-Egger |  | GSMR |  |
| --- | --- | --- | --- | --- | --- | --- | --- | --- | --- |
|  |  | OR | *P* | OR | *P* | OR | *P* | OR | *P* |
| T2D | Stroke | 1.11 | $1.33 \times 10^{-21}$ | 1.09 | $7.91 \times 10^{-09}$ | 1.09 | $7.87 \times 10^{-05}$ | 1.22 | $1.27 \times 10^{-43}$ |
| Stroke | T2D | 1.190 | $6.6 \times 10^{-3}$ | 1.09 | 0.11 | 1.349 | 0.35 | 1.181 | $6.2 \times 10^{-6}$ |

In the reverse direction, we have selected 23 IVs (sTable 7) for exposure (stroke) to determine the potential causal association between stroke and T2D. Though the IVW method suggested a modest significant causal association of stroke on T2D (OR = 1.19, 95% CI: 1.06–1.32, P = 6.6 × 10−³), but this finding was not supported by sensitivity analyses, including weighted median (OR = 1.09, 95%CI: 0.981-1.202, *P* = 0.11) and MR-egger OR = 1.349, 95% CI: 0.722-1.977, *P* = 0.35), indicating that the reverse causal relationship remains uncertain (Table 2).

### After conditional analysis MR results

Given that type 2 diabetes (T2D) is an established risk factor for coronary artery disease (CAD) and frequently co-occurs with it, we employed multi-trait conditional and joint analysis (mtCOJO) to condition T2D GWAS summary statistics on CAD, thereby mitigating its confounding genetic effects, prior to conducting two-sample Mendelian randomization (2SMR) analyses. From the conditioned T2D data, we identified 443 independent genome-wide significant SNPs (p< 5×10^−8^, LD r^2^<0.001) as instrumental variables (IVs) (sTable 8) to assess the causal effect of T2D (exposure) on stroke (outcome). In MR analysis, using the inverse variance weighted (IVW) method revealed a significant causal association (OR = 1.09, 95% CI: 1.07–1.12, P = 4.4 × 10^−23^). Sensitivity analyses, including weighted median (OR = 1.09, 95% CI: 1.03-1.103, *P* = 9.71×10^-06^), MR-Egger (OR= 1.09, 95% CI: 1.04-1.14, *P* = 6.7 x 10^-5^) also showed significant causal role of T2D on stroke risk (Table 3).

**Table 3.** Bidirectional causal genetic relationship between type 2 diabetes and stroke using MR analysis after considering confounder factors CAD.

| Exposure | Outcome | IVW |  | Weighted median |  | MR-Egger |  | GSMR |  |
| --- | --- | --- | --- | --- | --- | --- | --- | --- | --- |
|  |  | OR | *P* | OR | *P* | OR | *P* | OR | *P* |
| T2D | Stroke | 1.09 | $4.47 \times 10^{-23}$ | 1.07 | $9.71 \times 10^{-06}$ | 1.09 | $6.76 \times 10^{-05}$ | 1.09 | $9.42 \times 10^{-37}$ |
| Stroke | T2D | 1.14 | $4.3 \times 10^{-5}$ | 1.08 | 0.13 | 1.22 | 0.36 | 1.09 | $7.7 \times 10^{-6}$ |

In the reverse direction, we investigated the potential causal effect of stroke (exposure) on T2D (outcome) by conditioning stroke GWAS summary statistics on CAD using mtCOJO to address its confounding effects. From the conditioned stroke GWAs, we identified 20 independent genome-wide significant SNPs (p< 5×10^−8^, LD r^2^<0.001) as instrumental variables (IVs) (sTable 4.9) to evaluate the causal effects of stroke on T2D. The initial analysis with the IVW method indicated a causal association. However, sensitivity analyses using weighted median and MR-Egger methods failed to support a significant causal role of stroke on T2D risk (Table 3), suggesting inconsistency in the reverse direction.

Moreover, we also extended the conditioning of T2D to include CAD, body mass index (BMI), and educational attainment (EA) using mtCOJO, resulting in 625 IVs (stable 4.10). The IVW method in this refined analysis confirmed a robust causal effect of T2D on stroke risk (OR = 1.10, 95% CI: 1.08-1.12, *P* = 3.42 x 10^-26^), with consistent results across sensitivity analyses, weighted median (OR = 1.074, 95% CI: 1.04-1.103, *P* = 5.48 x 10^-08^), MR-Egger (OR= 1.06, 95% CI: 1.02-1.107, *P* = 1.95 x 10^-3^), MR-PRESSO (*P* = 6.42 × 10^-25^), and GSMR (*P* = 7.29 ×10^-30^) (Table 4). These findings provide substantial evidence of a causal role of T2D in elevating stroke risk, even after accounting for multiple confounders.

**Table 4.** Bidirectional causal genetic relationship between type 2 diabetes and stroke using MR analysis after considering confounder factors (BMI, CAD and EA).

| Exposure | Outcome | IVW |  | Weighted median |  | MR-Egger |  | GSMR |  |
| --- | --- | --- | --- | --- | --- | --- | --- | --- | --- |
|  |  | OR | *P* | OR | *P* | OR | *P* | OR | *P* |
| T2D | Stroke | 1.10 | $3.42 \times 10^{-26}$ | 1.074 | $5.48 \times 10^{-08}$ | 1.064 | $1.95 \times 10^{-03}$ | 1.085 | $7.29 \times 10^{-30}$ |
| Stroke | T2D | 1.21 | $1.45 \times 10^{-5}$ | 1.2 | $1.45 \times 10^{-4}$ | 1.45 | $5.92 \times 10^{-2}$ | 1.088 | $3.26 \times 10^{-5}$ |

OR: odds ratio; *P*: *P*-value; IVW: inverse variance weighted; MR: Mendelian randomization; MR-PRESSO: Mendelian Randomization Pleiotropy Residual Sum and Outlier.

Next, we repeated the above MR analyses considering stroke as exposure and T2D as the outcome. Here, we conditioned stroke on CAD, BMI, and EA, yielding 24 SNPs as IVs (p< 5×10^−8^, LD r^2^<0.001) for 2SMR analysis (sTable 11). The IVW method demonstrated a significant causal effect of stroke on T2D (OR = 1.21, 95% CI: 1.11-1.32, *P* = 1.45 x 10^-5^). Sensitivity analyses provided partial causality, with weighted median (OR = 1.2, 95% CI: 1.09-1.32, *P* = 1.89 x 10^-04^) and MR-PRESSO (*P* = 9.9 × 10^-04^). However, MR-Egger did not support a significant causal role of stroke on T2D (OR= 1.45, 95% CI: 1.007-2.09, *P* = 5.92 x 10^-2^) (Table 4), indicating potential pleiotropy or lack of robustness in this reverse relationship.

Overall, our MR analyses provide robust evidence for a putative causal effect of T2D on stroke risk, with consistent findings across multiple methods and confounder adjustments. In contrast, the reverse analyses, the initial evidence for stroke influencing T2D risk was weak and inconclusive, however, after conditioning on CAD, BMI, and EA, where IVW, weighted median, and MR-PRESSO align in detecting a significant effect. Notably, MR-Egger showed the association of stroke on T2D was not significant, suggesting the reverse causal relationship is weak and inconclusive compared to the forward direction.

### Identification of shared risk genes and pathways

To identify risk genes shared between T2D and stroke, we performed an SMR analysis using GWAS summary statistics for MTAG-boosted T2D and stroke GWAS data, respectively, with eQTLs datasets. SMR analysis revealed 109 for T2D and 25 genes for stroke (Bonferroni corrected *P-value* < 5.02 × 10^−6^, HEIDI P-value > 0.01, sTable 12-sTable 13). Notably, seven genes (*SREBF1, LTBP3, FAM234A, PABPC4, RMC1, KCNJ11, PLEKHA1*) overlapped between T2D and stroke, where 5 of them were novel for stroke (Table 5). Next, functional enrichment analysis of overlapping genes demonstrated enriched in pathways, including “positive regulation of cholesterol biosynthetic process”, “focal adhesion”, “Phosphatidylinositol 3-kinase signaling”, “regulation of autophagy”, “negative regulation of insulin secretion”, among others (Table 6).

**Table 5.** Identification of shared putative causal genes between type 2 diabetes and stroke.

| **Genes** | **CHR** | **topSNP** | ***p*_SMR_T2D** | ***P*_HEIDI_T2D** | ***p*_SMR_stroke** | ***p*_HEIDI_stroke** | **T2D** | **Stroke** |
| --- | --- | --- | --- | --- | --- | --- | --- | --- |
| *SREBF1* | 17 | rs9891957 | 8.91E-11 | 0.089 | 7.40E-09 | 0.329 | Known | Novel |
| *LTBP3* | 11 | rs12789028 | 1.04E-18 | 0.086 | 7.87E-09 | 0.318 | Known | Novel |
| *FAM234A* | 16 | rs113345834 | 1.04E-20 | 0.264 | 2.42E-09 | 0.518 | Known | Novel |
| *PABPC4* | 1 | rs3768321 | 2.19E-31 | 0.040 | 4.21E-08 | 0.833 | Known | Novel |
| *RMC1* | 18 | rs891389 | 9.18E-14 | 0.537 | 9.71E-08 | 0.973 | Known | Novel |
| KCNJ11 | 11 | rs2074310 | 2.51E-23 | 0.025 | 2.59E-12 | 0.464 | Known | Known |
| PLEKHA1 | 10 | rs11200608 | 6.32E-17 | 0.0128 | 1.26E-09 | 0.069 | Known | Known |

**Table 6.** Functional enrichment analysis of common genes shared between T2D and stroke.

| Pathways | GO Terms | Adjusted *P*-values | Odds Ratio | Combined Score | Genes |
| --- | --- | --- | --- | --- | --- |
| Positive regulation of cholesterol biosynthetic process | GO:0045542 | 0.0239 | 832.875 | 5287.722 | SREBF1 |
| Regulation of autophagy | GO:0010506 | 0.0239 | 33.06 | 192.974 | SREBF1, RMC1 |
| Negative regulation of insulin secretion | GO:0046676 | 0.024 | 208.09 | 1066.85 | KCNJ11 |
| Phosphatidylinositol 3-kinase signaling | GO:0014065 | 0.030 | 107.32 | 482.57 | PLEKHA1 |
| SREBP signaling pathway | GO:0032933 | 0.023 | 555.19 | 3338.1535 | SREBF1 |
| Regulation Of Cellular Catabolic Process | GO:0031329 | 0.034 | 53.577 | 204.86 | RMC1 |
| Insulin resistance | HP:0000855 | 0.018 | 133.12 | 626.09 | KCNJ11 |

## Discussion

The present study has comprehensively investigated the shared genetic relationships, molecular genetic factors, and pathways underlying type 2 diabetes and stroke by leveraging the largest GWAS datasets. Our analyses have disclosed significant genetic overlap, genetic correlation, and a putative causal relationship between T2D and stroke, suggesting that genetic elements contribute to T2D and stroke comorbidity. Cross-disorder meta-analysis has identified 10 novel lead-independent SNPs shared between T2D and stroke, which were not previously identified as genome-wide significant in individual GWAS studies. The gene-based study has led to prioritizing five novel risk genes for stroke (Table 5).

The gaussian causal mixer model method has suggested substantial genetic overlap exists between T2D and stroke and significant positive global genetic correlations. These findings indicate that shared genetic factors drive the association between T2D and stroke. This finding is consistent with previous studies, although their T2D datasets were not as large as the one used in this study [31–33]. Next, we estimated the causal effect of the genetic factors on stroke using an MR-based approach. To eliminate the obscuring effects of putative comorbid conditions and factors, we have done conditional analysis to capture the specific contribution of genetic factors of each trait using mtCOJO followed by MR analysis. This approach provides a more refined understanding of the causal relationship between T2D and stroke risk.

We used MTAG, a most powerful meta-analysis method that improves the statistical power of the individual GWAS datasets to identify trait-specific genetic associations of T2D and stroke. Utilizing MTAG, GWAS datasets for T2D and stroke were used as an input in FUMA, we identified 10 novel independent genome-wide significance SNPs shared between T2D and stroke (Supplementary Table S6); those SNP were also colocalized with both T2D and stroke, suggesting that genetic variations influence disease risk. Among the identified novel independent lead SNPs, rs35801464, rs10885414, and rs116369954, which encode the *TCF7L2* gene associated with regulating plasma glucagon level [34]. Previously, the *TCF7L2* gene was reported to be involved in developing T2D in multiple ethnicities [34].

Our SMR analysis integrating GWAS with eQTLs datasets identified putative causal risk genes associated with T2D and stroke, respectively. Among seven common genes (*SREBF1, LTBP3, FAM234A, PABPC4, RMC1, KCNJ11, PLEKHA1*) shared between T2D and stroke. *SREBF1* encodes sterol regulatory element binding transcription factor 1 protein, which plays an important role in the regulation of fatty acid, triglyceride, and cholesterol synthesis and is associated with obesity, type 2 diabetes risk, and dyslipidemia [35–38]. Dyslipidaemia or hypercholesterolemia is a well-established risk factor for atherosclerosis and plays a significant role in the progression of ischemic stroke [39]. The novel independent lead SNP, rs4925118, within *SREBF-1,* may have a causal role in T2D and stroke risk. *LTBP3* encodes for latent transforming growth factor beta-binding protein 3. This protein helps control islet cells, which keeps glucose levels stable [40]. Mutations in the *LTBP3* gene have been associated with thoracic aortic aneurysm and dissection (TAAD), and patients with TAAD have a higher risk of developing ischemic and hemorrhagic strokes [41].

Moreover, pathway-based functional enrichment analyses of common causal genes revealed that those genes are associated with regulating cholesterol biosynthetic processes. Patients with T2D often exhibit lipid abnormalities, such as increased cholesterol and fatty acid accumulation in pancreatic β-cell, leading to pancreatic islet degeneration. Overexpression of SREBP-2 in β-cells leads to cholesterol buildup, impairing cell function, reducing insulin secretion, and contributing to T2D [42]. Additionally, cholesterol homeostasis is crucial in the pathogenesis of several diseases, including cardiovascular diseases (CVD) and neurological disorders [43]. In stroke, high levels of LDL cholesterol increase the risk of ischemic stroke by promoting atherosclerosis, while low levels raise the risk of intracerebral hemorrhage [44]. We also found that causal genes are enriched in the phosphoinositide 3-kinases (PI3Ks) pathway, associated with glucose metabolism. Dysregulation of the PI3K pathway is associated with insulin resistance and ultimately leads to T2D development [45]. PI3K controls how endothelial cells and neutrophils interact during ischaemic injury, a condition where oxidative stress, neutrophil adhesion, and trans-endothelial migration play a major role. Moreover, PI3K is a critical signalling pathway that controls cell migration, proliferation, differentiation, and apoptosis. It also plays a crucial role in oxidative stress development and the progression of stroke pathogenesis [45]. Therefore, the common causal genes and biological processes could be a potential target for developing diagnostics and therapeutic approaches for both T2D and stroke.

While the key strengths of this study that it provides robust evidence of putative causal association of T2D on the risk of stroke. The causal effect signals of T2D on stroke was not attenuated significantly despite conditioning T2D on CAD suggest this signal is unlikely confounded or mediated through CAD. In addition, cross-trait meta-analysis and functional annotations of the SNPs via causal gene-based methods identified risk genes and pathways that underlie shared architecture between T2D and stroke. However, our study has several limitations. Firstly, despite the substantial polygenicity of both T2D and stroke, we noted the relatively small sample size of stroke GWAS may have limited power in identifying causal effect of stroke on T2D, although current results do not strongly support this causality.

### Conclusions

In summary, the findings of this study suggest significant shared genetic architecture between T2D and stroke, and T2D showed robust evidence of causal relationship of T2D on stroke risk. Our cross-trait meta-analysis identified novel independent lead SNPs shared between T2D and stroke. The SMR analysis has suggested that pleiotropic and/or causal genes are shared between T2D and stroke. The shared biological pathways enriched by pleiotropic genes were also identified. The findings advance the current understanding of genetic aetiology and biological mechanisms, which could facilitate the development of new therapeutic approaches and points of intervention for these co-occurring diseases.

## Data Availability

All data used in this manuscript are publicly available.

## Data Availability Statement

All data here analyzed is publicly available on GIGASTROKE consortium and the DIAGRAM consortium.

## Acknowledgments

We thank the GIGASTROKE and DIAGRAM project for making stroke and typ2 diabetes GWAS summary statistics data publicly available. I would like to acknowledge Research Training scholarship from The University of Queensland, Australia.

## Author contributions

TI, JZ, MAM contributed to the study conception, design critical reading and editing. TI performed bioinformatics analysis, prepared the figures and wrote the manuscript. All authors read and approved the final manuscript.

## Conflict of Interest

“Authors declare no conflict of interest”.

**Supplementary Tables (1-13).**

